# Optimal Control Measures to Combat COVID 19 Spread in Sri Lanka: A Mathematical Model Considering the Heterogeneity of Cases

**DOI:** 10.1101/2020.06.04.20122382

**Authors:** WPTM Wickramaarachchi, SSN Perera

## Abstract

The COVID 19 pandemic caused by the novel corona virus (SARS-CoV-2) has been one of the major public health concerns across the globe, currently more than 3.5 million individuals have been infected, and the number of deaths has passed 250,000. The world wide burden of the disease has been massive, and the governments are in dilemma to protect the health system of the country while safeguarding the economy. There is no vaccine or antivirus drug found against this virus while multiple research groups are actively working on a suitable candidate. The only available mode of minimizing the disease burden has been to control its transmission among the population. Since the occurrence of first COVID 19 local case on 11 March 2020, the government of Sri Lanka introduced serious social distancing and public health interventions in its fullest capacity as a developing nation to effectively combat with the disease spread. This study focuses to develop a mathematical model to investigate the dynamic of this novel disease using an extended version of an SEIR compartmental structure considering the heterogeneity of cases such as asymptomatic, symptomatic with mild indications and the cases required intensive care treatments. All the measures and interventions are in progress with a significantly large social and economic cost, thus, optimal control techniques are used to identify the most appropriate strategies to minimize this cost. The results of the simulations prove that optimal control measures can be worked out as the epidemic curves are flattened while delaying the outbreak so that the health system might not be under pressure to treat and care the patients.

## 1 Introduction

The COVID 19 outbreak occurred in the city of Wuhan, Hubei province, China during late December 2019 from a cluster of pneumonia cases. The Chinese health authorities identified and informed World Health Organization (WHO) that the pneumonia condition was due novel beta corona virus, the 2019 novel virus (2019-nCoV, recently renamed as SARS-CoV-2, the cause of corona virus disease COVID-19) [1]. It is claimed that the novel corona virus likely to have originated from a zoonetic type of transmission, occurred in a wet sea food market where wild animals are sold openly. Soon after few days, Chinese researchers found out that the corona virus effectively show the human-to human transmission, and this new virus was identified to be extremely contagious among people [2].

The novel corona virus transmitted to human through respiratory droplets of another. It had also been revealed later that these droplets can survive in variety of surfaces for multiple hours or even days. Common symptoms of COVID 19 disease have been fever, cough and fatigue. There are some less common symptoms including sputum production, headache, hemophiliacs, and diarrhea [3]. According to WHO, COVID 19 has spread for more than 210 countries and independent territories while Italy, Spain, United States and Iran are the hardest hit apart from China where the disease is known to be emerged but now significantly controlled and stable. In numbers, currently more than 30 million people have been infected while there are 200 000 reported deaths worldwide [4].

Since this is a new virus, there is no vaccination found yet, however, there are number of experiments are in progress including animal and human trials, across the globe to find a successful vaccine candidate to fight with the corona virus [5]. Researchers claim that, though they are able to find a suitable vaccine, it would take reasonable number of months to make them available to people. The only effective strategy to combat with COVID 19 is to control its transmission through social distancing measures and public health interventions. Contact tracing and isolation of cases is a common intervention for controlling infectious disease outbreaks which most of the countries have been following, however, it might need intensive public health effort and community mobilization due to the requirement of figuring out all possible contacts. Current modeling outcomes suggest that at a minimum of 80% of symptomatic contacts must be traced, isolated and treat to maintain the efficacy of control and the stability of the disease spread [6].

In Sri Lanka, the first COVID 19 case was found on 26 January 2020, was a Chinese national and she recovered after few weeks. The first local patient was found on 11 March 2020 and the government of Sri Lanka took strong decisions to control the transmission of the disease over the community including shutting down all the places of public gatherings such as schools, universities and non essential services, imposing travel ban to high risk countries, introducing mandatory quarantine for all arrivals to the country, shutting down the air port and finally imposing island wide curfew [7]. The time line of COVID 19 related events and responses by the government is illustrated in Figure 1. As of 28 April 2020, there are 600 confirmed cases, 134 recoveries and 7 deaths reported in the island while there are many suspected exposed cases are closely monitored [7]. Few of the high risk areas and villages have been locked down restricting any type of mobility. Even though the public health sector including the military forces are acting effectively, one of the major challenges to combat with the virus in Sri Lanka has been the significant rise in the asymptomatic infections who are not showing any COVID 19 symptoms but they are carries of the virus in the population [8].

**Fig. 1:**
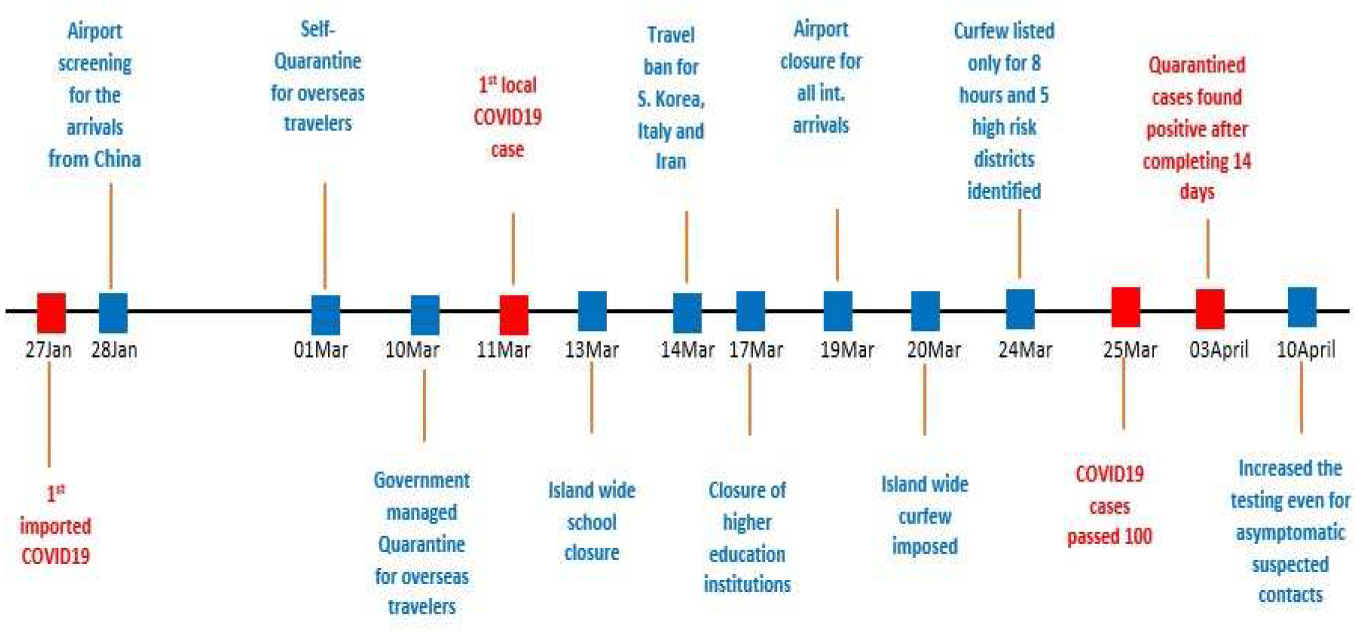
COVID19 events (red) and control measures (blue) in Sri Lanka from the first reported case.

Since COVID 19 is a new disease emerged in the world, lack of data is available related to the dynamics so that they fit to existing mathematical models to predict the outbreak. However, these models may be used to demonstrate the possible different scenarios of the disease transmission with respect to social distancing and public health intervention measures introduced by authorities [9].

In this study, we use the SEIR (Susceptible, Exposed, Infected, Recovered) compartmental approach to model the dynamic of COVID 19 in Sri Lanka. As it it critical to test, trace and isolate not only the symptomatic cases but also the asymptomatic, the infected population then is divided into asymptomatic (*I_A_*) and the symptomatic with mild symptoms (*I_M_*). In the context of Sri Lanka, all cases who are tested positive for the virus are isolated in designated hospitals and treated. The patients whose condition is not developed for the severe level (*I_H_*) are treated in isolated general wards however, the patients whose condition is worsen due to their demography and various other health related issues (*I_C_*) are transferred to Intensive Care Units (ICUs) [10]. In order to deal with this COVID 19 outbreak, the government has decided to implement tough measures such as social distancing, personal protection, aggressive testing for the virus of all contacts and etc. In this context, the optimal control problem is considered to study the effect of said control measures to spread the COVID 19 disease [11,12].

This manuscript is organized as follows: In section 2, the deterministic mathematical model without control is discussed. Furthermore, basic analysis and the disease free equilibrium are presented by defining the basic reproduction number (*R*_0_). In section 3, we present the optimal control problem with essential mathematical analysis. Numerical results and discussion are given in section 4 and finally the conclusion is presented in section 5.

## 2 Methods

### 2.1 Mathematical Model with out Control

First, we introduce the mathematical model of COVID19 transmission with out any control measures. A more extended version of the SEIR (Susceptible-Exposed-Infected-Recovered) compartment model structure is used to formulate this dynamic [10,13–15]. In Sri Lanka, the health authorities treated all the symptomatic COVID19 cases in government hospitals, rather than advising them to be self-isolated. However, recent international travelers and close contacts of the identified COVID19 patients are isolated in government managed quarantine centers in the different parts of the island [7]. If patients are identified from those groups then they are immediately taken to the hospitals and treated. However, it is also found that a reasonable number of individuals who are tested positive while they were asymptomatic [7,5]. Based on this policy structure in Sri Lankan context, seven population compartments are considered for the model; Susceptible (*S*), Exposed (*E*), Infected with asymptomatic (*I_A_*), Infected with mild symptoms (*I_M_*), Isolated in designated hospitals (*I_H_*), Patients with critical conditions treated in Intensive Care Units (*I_C_*) and the patients who clinically determined as Recovered (*R*) [10]. Following the compartmental transition schematic diagram illustrated in Figure 2, the seven dimensional differential system describing the COVID19 transmission is given by

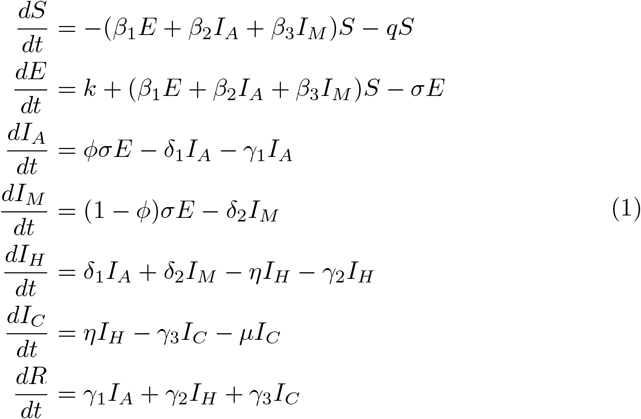

where *β*_1_, *β*_2_ and *β*_3_ represent the transmission rates from the exposed, infected and asymptomatic, and infected and symptomatic respectively while *q* is the rate of isolation of the susceptible individuals due to lock down, *k* is the rate of imported exposed cases, *σ* is the rate at which the exposed cases become infected, *φ* is the percentage of exposed individuals who become asymptomatic, *δ*_1_ is the rate at which the asymptomatic cases are tested and hospitalized, *δ*_2_ is the rate at which the symptomatic cases are tested and admitted to hospitals, *η* is the rate of patients condition becomes severe and require intensive care treatments, *γ*_1_ is the recovery rate of asymptomatic cases who are not in hospitals, *γ*_2_ is the recovery rate of mild symptomatic cases who are in general wards in hospitals, *γ*_3_ is the recovery rate of critically sick patients and *µ* is the death rate of the disease.

The initial conditions for the model (1) is as *S*(0) = *S*^0^, *E*(0) = *E*^0^, *I_A_*(0) = 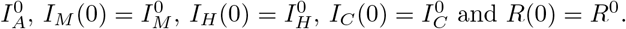 and *R*(0) = *R*^0^.

**Fig. 2:**
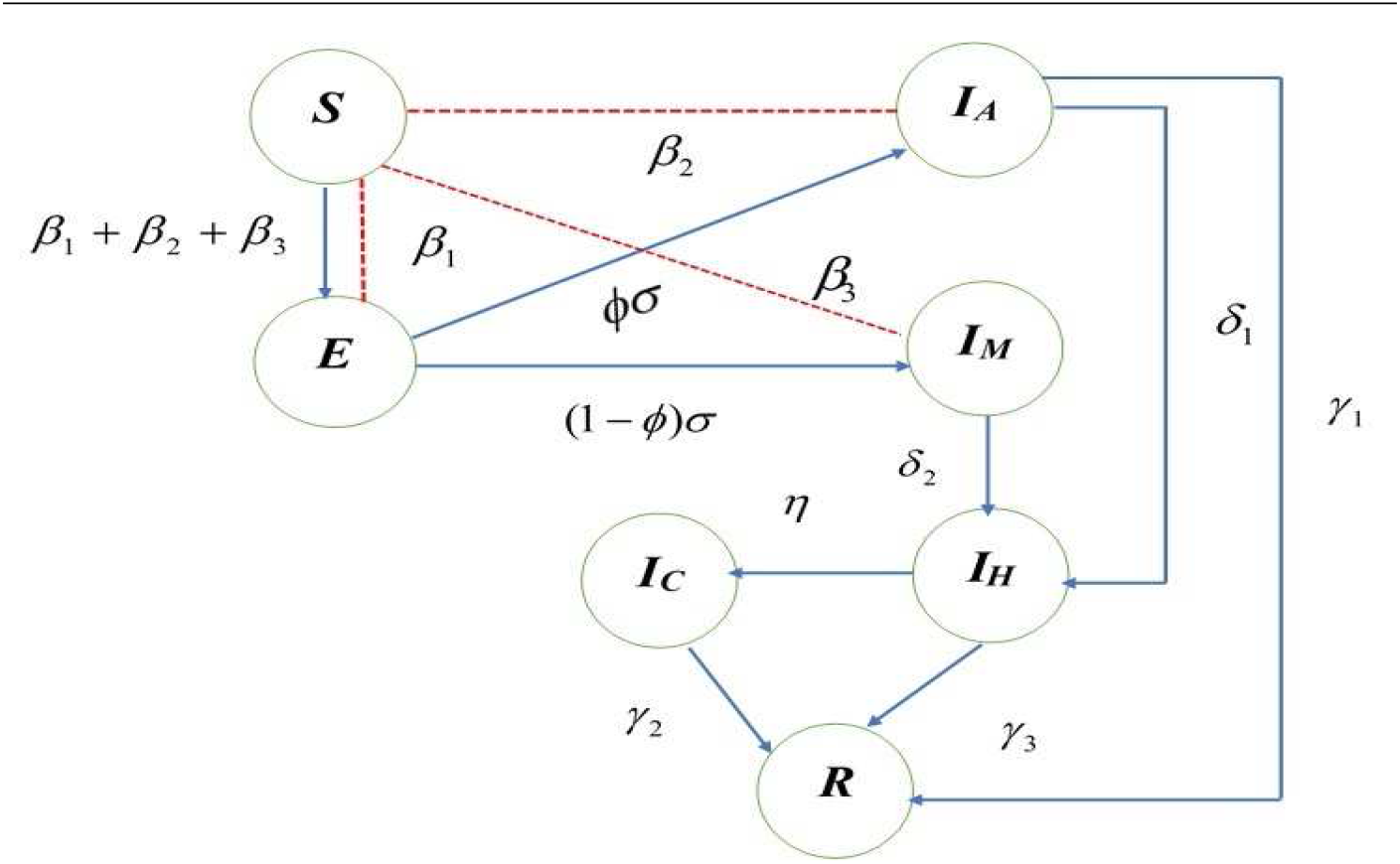
Schematic diagram of COVID19 transmission

We let the set of solutions denoted by *Ω* to the system of nonlinear differential equations in (1) as

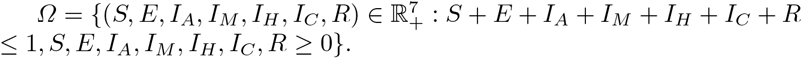

### 2.2 Analysis of the Model

#### 2.2.1 Basic Reproduction Number

Basic reproduction number *R*_0_ stands for the number of secondary infections those can be produced by a single infected patients on average [18]. It is very critical to distinguish new infections in the dynamic of the population to compute *R*_0_. In general, we let *x* = (*x*_1_*,…, x_n_*)^*T*^, *x_i_ ≥* 0, be the number of individuals in each population class. For simplicity, we arrange the compartments in such a way that first *m* stand for the infected individuals. We also define the set X_0_ = {*x ≥|x_i_* = 0, *i* = 1,…, *m*}.

Let *ℱ_i_*(*x*) be the rate of arrival of new infections in compartment 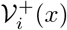 be the rate of transfer of individuals into compartment *i* in various other routes, and 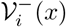 be the rate of transfer of individuals out of compartment *i*. The functions 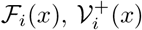 and 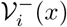 are assumed to be continuous and at a minimum of twice differentiable on *x*. Now in general terms, the system of differential equations can be represented in the form

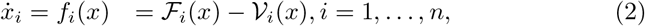

where 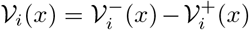 and the above functions must meet the assumptions A(1)-A(5) listed below.

A(1) Since each function represents a directed transfer of individuals in the population, they are all non-negative. That is, if *x ≥* 0, then 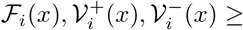 0 for *i* = 1*,…, n*.
A(2) If a compartment is empty, then there can be no transfer of individuals out of the compartment by death, migration, infection, nor any other means. That is, if *x_i_* = 0 then 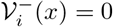.
A(3) The incidence of infection for uninfected compartments is zero. That is, ℱ_*i*_(*x*) = 0 if *i > m*.
A(4) If the population is free of disease then the population will remain free of disease. Thus, if *x ∈* **X**_0_ then *ℱ_i_*(*x*) = 0 and 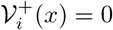 for *i,…, m*.
A(5) If the population is held closed to the Disease Free Equilibrium (DFE) then the population will get back to the DFE as ruled by the linearized system

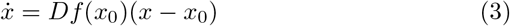

where 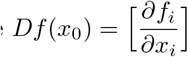 assessed at the DFE, *x*_0_. This can be written as if *ℱ*(*x*) = 0 then all eigenvalues of *Df*(*x*_0_) have negative real parts.

Using th assumptions A(1)-A(5) enable us to partition the matrix *Df*(*x*_0_). This is given by the following lemma.

**Lemma 1** *If x*_0_ *is a DFE of the system (2) and f_i_*(*x*) *satisfies A(1)-A(5) then the derivatives DF*(*x*_0_) *and DV*(*x*_0_) *are partitioned as*

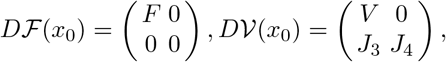

*where F and V are the m × m matrices defined by* 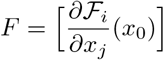 and 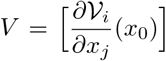 *with* 1 *≤ i, j ≤ m. Further, F is non-negative, V is a non-singular M-matrix and all eigenvalues of J*_4_ *have positive real part*.

**Proof** Let *x*_0_ *∈* **X**_0_ be a DFE. By A(3) and A(4), 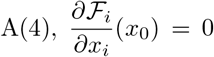 if either *i > m* or *j > m*. Similarly, A(2) and A(4) gives that if *x ∈* **X**_0_ then *𝒱_i_*(*x*) = 0 for *i ≤ m*. This provides 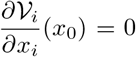 for *i ≤ m* and *j > m*. This shows the stated partition and zero blocks. The non-negativity of *F* follows from A(1) and A(4).

Let *e_j_* be the Euclidean basis vectors. That is, *e_j_* is the *j*th column of the *n × n* identity matrix. Then, for *i* = 1*,…, m*

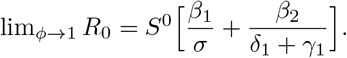

To show that *V* is a non-singular M-matrix, note that if *x*_0_ is a DFE, then using A(2) and A(4), *𝒱_i_*(*x*_0_) = 0 for *i* = 1*,…, m* and if *i* ≠ *j*, the the jth component of *x*_0_ + *he_j_* = 0 and *𝒱_i_*(*x*_0_ + *he_j_*) *≤* 0, by A(1) and A(2). Therefore, 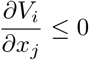 for *i ≤ m* and *j* ≠ *i* and *V* has the *Z* sign pattern [16]. Furthermore, by A(5), all eigenvalues of *V* have positive real parts. These two conditions provide that *V* is a non-singular M-matrix [16]. Finally, A(5) also implies that the eigenvalues of *J*_4_ have positive real part. This completes the proof.

Now we aim to compute the basic reproduction number for the system (1). The method of next generation matrix is used to derive *R*_0_. For this purpose we now define the new vector of only infected variables *<u>X</u>* = (*E, I_A_, I_M_*) containing the classes which are responsible to transmit the virus in the population. It is assumed that the classes of *I_H_* and *I_C_* are fully isolated and it is unlikely that the virus is transmitted to the society anymore. Hence, we establish the following system of differential equations [16–18]:

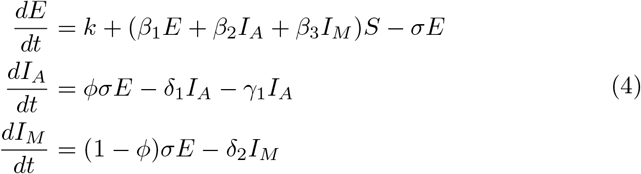

To apply the next generation matrix method, the necessary matrices *F* and *V* are obtained as follows [16,17]:

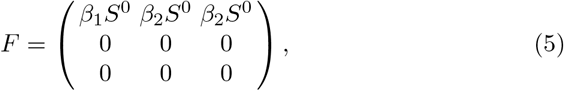

and

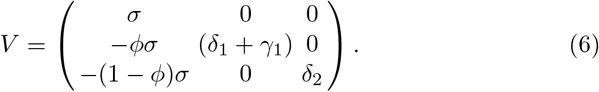

Now the next generation matrix system is

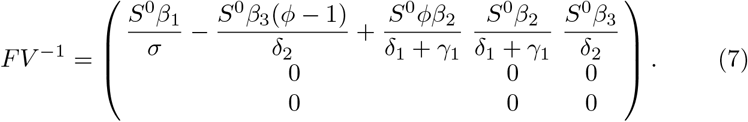

So, the basic reproduction number is the spectral radius *ρ* of the matrix *F V*^−1^. Thus, we obtain

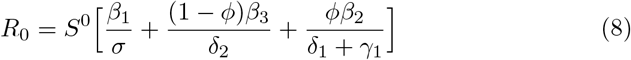

The expression for *R*_0_ reveals very useful information about the dynamic of COVID19 transmission such that the expected number of secondary infection is the addition of infections due to the exposed, asymptomatic, symptomatic cases respectively. As *φ* goes to 1, the secondary infections are not produced by the cases with mild symptoms as they have been tested and isolated early. Mathematically, it can be very easily shown that

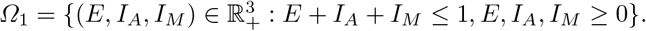

#### 2.2.2 Stability Analysis of the Disease Free Equilibrium

Let us first obtain matrix *M* such that

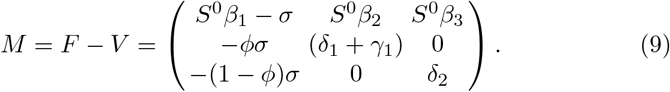

Now define *s*(*M*) = max*{Re*(*α*): *α* is an eigenvalue of *M}*. Note that *s*(*M*) is a simple eigenvalue of *M* with a positive eigenvector. In relation to *R*_0_ we can establish the following equivalences: *R*_0_ *>* 1 if and only if *s*(*M*) *>* 0 and *R*_0_ < 1 if and only if *s*(*M*) < 0.

Let us now define the set of solution to system (4) by

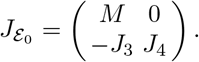

**Theorem 1** *If R*_0_ < 1 *then the DFE, ε*_0_ *is locally asymptotically stable on Ω*_1_.

**Proof.** To prove this we need to apply the assumptions A(1)-A(5) and A(1)-A(4) are easily verified. For A(5) we need to show that the matrix

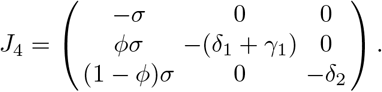

have negative real parts with *J*_3_ = *−F*,

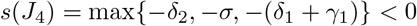

We then compute the eigenvalues of *J*_4_ and yield,

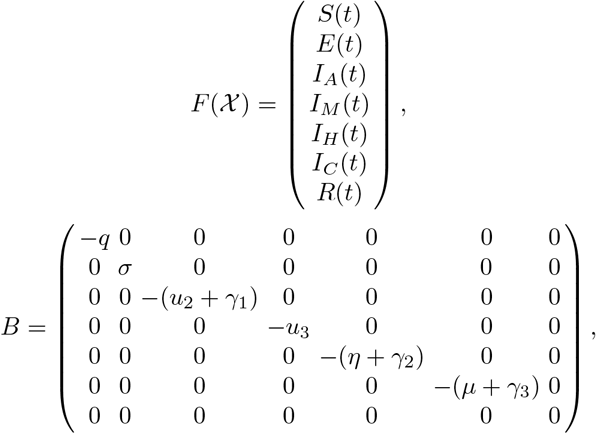

Thus, if *R*_0_ < 1 then the DFE, *E*_0_ is locally asymptotically stable.

## 3 Optimal Control

It is very clear that the only available strategy to combat with the novel corona virus is to control its spread over the population. Controlling can be achieved by reducing the transmission rates [19]. In our model in system (1), the spread of the virus is mainly due to three population compartments, exposed, infected with asymptomatic and infected with mild symptoms, and non of the three groups are isolated until the individuals are being clinically tested. The asymptomatic cases have been a very serious concern for the public health system across the globe including Sri Lanka. It has been estimated that around 20% of the cases may be asymptomatic hence they are undetected, however, with the potential of spreading the virus over the population. In this section, we introduce control measures to the system (1) and modify our model and necessary mathematical derivations and analysis will be carried out.

### 3.1 Mathematical Model with Control

In the model with control, we introduce the combined factor (1*−u*_1_) to reduce the transmission rates *β*_1_, *β*_2_ and *β*_3_ respectively from exposed, infected with asymptomatic and infected with mild symptoms population classes. Thus, this *u*_1_ measures the effort of personal protection such as wearing face marks, personal hygiene practices, social distancing methods and etc. The control variable *u*_2_ measures the rate of identifying asymptomatic cases through contact tracing, testing and isolating them to treat in designated hospitals. The control variable *u*_3_ measures the rate of tracing, testing and isolating of patients with mild symptoms. In this model, we assume that *u*_2_*I_A_* and *u*_3_*I_M_* are removed from *I_A_* and *I_M_* compartments and they are added to the compartment *I_H_*. In addition, the critically sick patients who are currently at *I_H_* compartment will be transferred to the class of patients in intensive care units with a rate of *η*. It is further assumed that asymptomatic cases who are undetected and could be recovered themselves with a rate of *γ*_1_, patients who are in general wards with mild symptoms are recovered with a rate of *γ*_2_ and the patients in ICUs are recovered with a rate of *γ*_3_, and all are added to the recovery compartment. The modified version of the system (1) can now be established as in system (10).

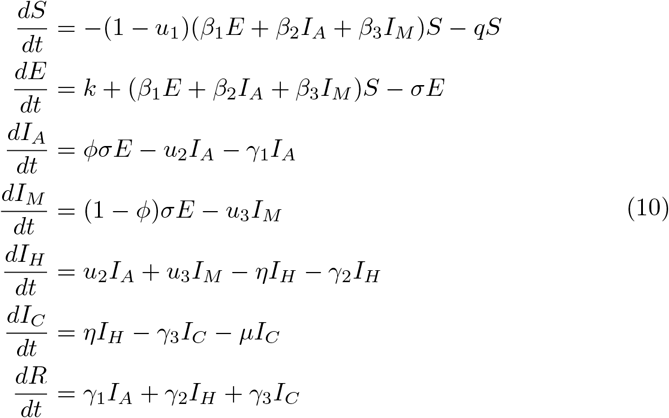

### 3.2 Mathematical Analysis of the Model with Control

It is clear that we have introduced three time invariant control variables *u*(*t*) = (*u*_1_*, u*_2_*, u*_3_) *∈ 𝒰* into system (1) and these variables are associated with the population compartments *S, E, I_A_, I_M_* and *I_H_*. Further, the control variables are bounded and measurable such that

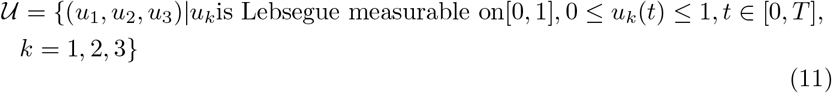

The objective functional for the control problem in (10) is now defined as

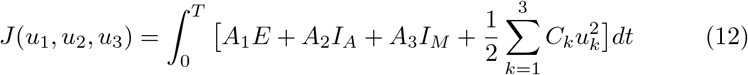

subject to (10).

It is aimed to minimize the cost functional in (12) which consists of populations exposed (*E*), asymptomatic infected (*I_A_*) and mildly infected (*I_M_*) as well as the socio-economic cost related to wearing masks, sanitizing methods, cost of social distancing measures, and etc given by 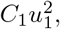, public health cost on contact tracing, testing and isolation of asymptomatic cases given by 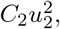, and the same cost that is for cases with mild symptoms represented by 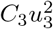. The constants *A*_1_, *A*_2_, *A*_3_, *C*_1_, *C*_2_ and *C*_3_ are the weights and balancing parameters and they measure the associated relative cost of the interventions over the interval [0*, T*]. We find the optimal control measures 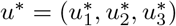 such that

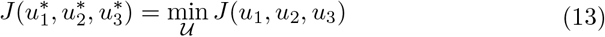

Now we derive necessary conditions to find the solution for the optimal control problem using Pontryagins Maximum Principle [17,19,21,22]. to show the existence of the control problem, we rewrite the system (10) as in the following form [17,20].

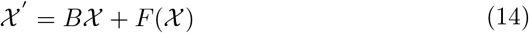

where

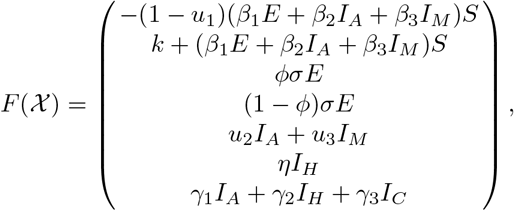

and

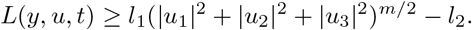

and 𝒳′ is the derivative of *𝒳* with respect to time.

To show the uniform Lipschitz continuity, we let

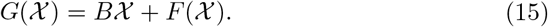

The function *F* (*𝒳*) in equation (15) satisfies *|F* (*𝒳*_1_) *− F* (*𝒳*_2_)*| ≤ Z*_1_*|S*_1_ *− S*_2_*|* + *Z*_2_*|E*_1_ *− E*_2_*|* + *Z*_3_*|I_A_*_1_ *− I_A_*_2_*|* + *Z*_4_*|I_M_*_1_ *− I_M_*_2_*|* + *Z*_5_*|I_H_*_1_ *− I_H_*_2_*|* + *Z*_6_*|I_C_*_1_ *− I_C_*_2_*|* + *Z*_7_*|R*_1_ *− R*_2_*|*.

Now choose *Z >* 0 such that *Z* = max(*Z*_1_*, Z*_2_*, Z*_3_*, Z*_4_*, Z*_5_*, Z*_6_*, Z*_7_). Thus, we have

*|F* (*𝒳*_1_) *− F* (*𝒳*_2_)*| ≤ Z*(*|S*_1_ *− S*_2_*|* + *|E*_1_ *− E*_2_*|* + *|I_A_*_1_ *− I_A_*_2_*|*

+ *|I_M_*_1_ *− I_M_*_2_*|* + *|I_H_*_1_ *− I_H_*_2_*|* + *|I_C_*_1_ *− I_C_*_2_*|* + *|R*_1_ *− R*_2_*|*).

Further we have *|G*(*𝒳*_1_) *− G*(*𝒳*_2_)*| ≤ Z|𝓍*_1_ *− 𝓍*_2_*|* with *Z* = *Z*_1_ + *Z*_2_ + *Z*_3_ + *Z*_4_ + *Z*_5_ + *Z*_6_ + *Z*_7_ + *kKk < ∞*.

Therefore, the function *G*(*𝓍*) is uniformly Lipschitz continuous. Hence we can state that the solution of the control system in (10) exists.

**Theorem 2** *Given the objective functional J*(*u*_1_*, u*_2_*, u*_3_) *according to (12), where the control set 𝒰 given by (11) is measurable subject to (10) with initial condition for the problem at t* = 0*, then there exists an optimal control* 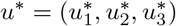 *such that* 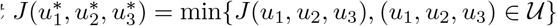

**Proof.** It is noted that the state variables and the control variables in the problem (10) are nonempty and the set *U* contains the control variables is closed and convex. The right hand side of system (10) is continuous, bounded above and can be written as a linear function of *u* with time invariant coefficients and are depending on state. There exist constants *l*_1_*, l*_2_ *>* 0 and *m >* 1 such that the intergrand *L*(*y, u, t*) of the objective functional *J* is convex and it satisfies

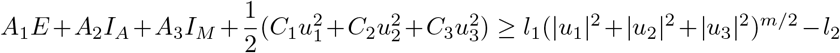

The state variables and the set of control *U* is clearly bounded and nonempty. The solutions are bounded, and convex. Thus, the system is bi-linear in control variables as the solutions are bounded. Now the following is verified so that

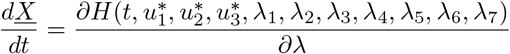

where *A*_1_*, A*_2_*, A*_3_*, C*_1_*, C*_2_*, C*_3_*, l*_1_*, l*_2_ *>* 0 and *m >* 1 [23,24].

Now we discuss the method of obtaining the solution to the problem (10). For this, it is necessary to define the Lagrangian and Hamiltonian for the optimal control problem (10). Thus, the Lagrangian *L* is stated as

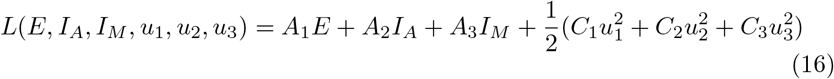

and for the Hamiltonian *H* we let *<u>X</u>* = (*S, E, I_A_, I_M_, I_H_, I_C_, R*), *𝒰* = (*u*_1_*, u*_2_*, u*_3_) and *λ* = (*λ*_1_*, λ*_2_*, λ*_3_*, λ*_4_*, λ*_5_*, λ*_6_*, λ*_7_), and we write

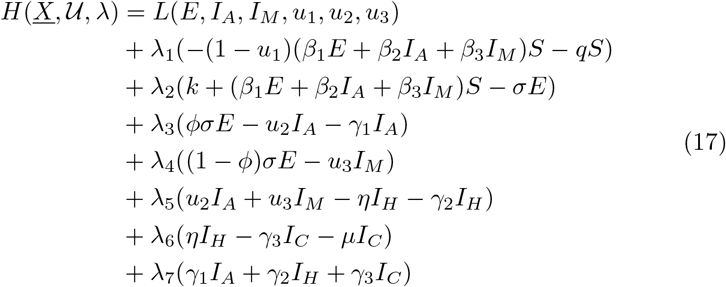

where λ_*j*_, *j* ∈ {*S*, *E*, *I_A_*, *I_M_*, *I_H_*, *I_C_*, *R*} are the adjoint variables. Next derivation is the necessary conditions for the Hamiltonian *H* given in (17).

**Theorem 3** *Given an optimal control* 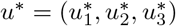 *and a solution* 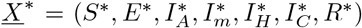 *with respect to the system (10), there exist adjoint variables λ_j_, j ∈ {S, E, I_A_, I_M_, I_H_, I_C_, R} satisfying*

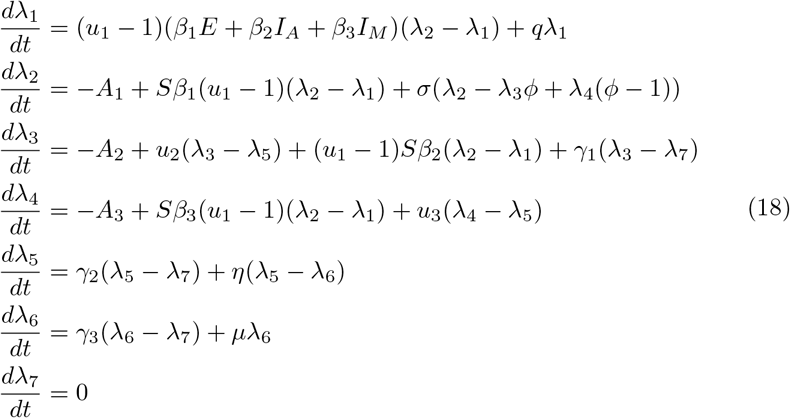

*with transversality conditions*

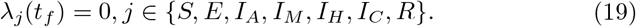

*In addition, the optimal control functions* 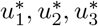 *are given by*

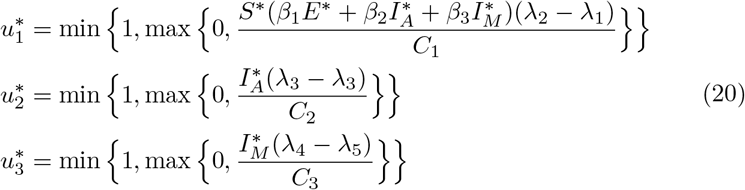

**Proof.** The control system (10) is obtained by taking the derivative

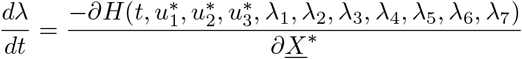

and the adjoint system (18) is obtained taking

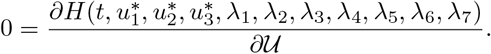

and the optimal control measures can be derived using

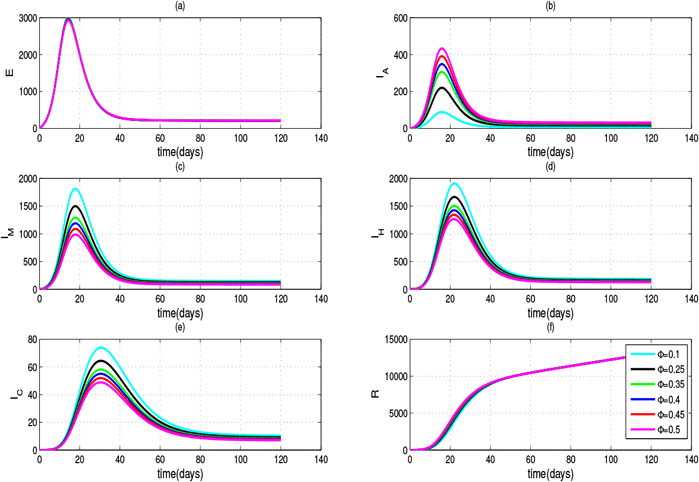

## 4 Numerical Results and Discussion

In this section, we obtain the numerical solutions for the problem with out control (1) and for the control problem (10). The Runge-Kutta algorithm of order four is implemented in MATLAB to solve the problem with out control and the numerical schemes presented in [25–27] are coupled with Runge-Kutta method of order four to carry out the simulation for the problem with control.

### 4.1 Algorithm for the Optimal Control Problem

STEP 0: Guess an initial estimation to control parameters *u* and *t_f_*.
STEP 1: Use initial conditions *S*(0)*, E*(0)*, I_A_*(0)*, I_M_*(0)*, I_H_*(0)*, I_C_*(0) and *R*(0) and the stocked values by *u* and *t_f_*. Find the optimal states 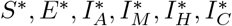 and *R^∗^* which iterate forward in the control problem (10)-(20).
STEP 2: Use the stocked values by *u* and the transversality conditions *λ_j_*(*t_f_*) for *j* = 1, 2, 3, 4, 5, 6, 7 while searching the constant *λ*_7_(*t_f_*) using the scant-method. Find the adjoint variables *λ_j_*(*t_f_*) for *j* = 1, 2, 3, 4, 5, 6, 7 which iterate backward in the control problem (10)-(20).
STEP 3: Update the control utilizing new state variables *S, E, I_A_, I_M_, I_H_, I_C_, R* and *λ_j_*(*t_f_*) for *j* = 1, 2, 3, 4, 5, 6, 7 in the characterization of optimal *u^∗^* given in (20).
STEP 4: Test the convergence. If the values of the sought variables in this iteration and the final iteration are sufficiently small, check out the recent values as solutions. If the values are not small, go back to STEP 1 [28–30].

### 4.2 Simulation of the COVID 19 Dynamic System with out Control

Figure 3 shows the simulation results of the problem with out control measures given in (1). It is found recently that there are a significant number of asymptomatic cases with in the populations who are also carriers of the virus. In the public health perspectives, it is very critical to clinically identify these cases through aggressive testing and isolate them if they are positive for the virus. The outcome of this task depends on how many cases are asymptomatic as a proportion. Therefore, we aim to assess the sensitivity of this proportion in the parameter level. Thus, we let *φ* to be varying and consider the vector of values *φ* = (0.1, 0.25, 0.35, 0.4, 0.45, 0.5) for this simulation. The rest of the parameters are *β*_1_ = 0.5*, β*_2_ = 0.6*, β*_3_ = 0.45*, γ*_1_ = 0.5*, γ*_2_ = 0.2*, γ*_3_ = 0.05*, δ*_1_ = 0.15*, δ*_2_ = 0.25*, η* = 0.005*, µ* = 0.04*, σ* = 1*/*5*, φ* = 0.25, *k* = 0.00405 and *q* = 0.0004. The initial conditions for the dimensionless form of the problem are *S*(0) = 0.85*, E*(0) = 0*, I_A_*(0) = 0*, I_M_*(0) = 0*, I_H_*(0) = 0*, I_C_*(0) = 0 and *R*(0) = 0 [10,13]. No control measures *u*_1_*, u*_2_ and *u*_3_ are inactive in this case.

**Fig. 3:**
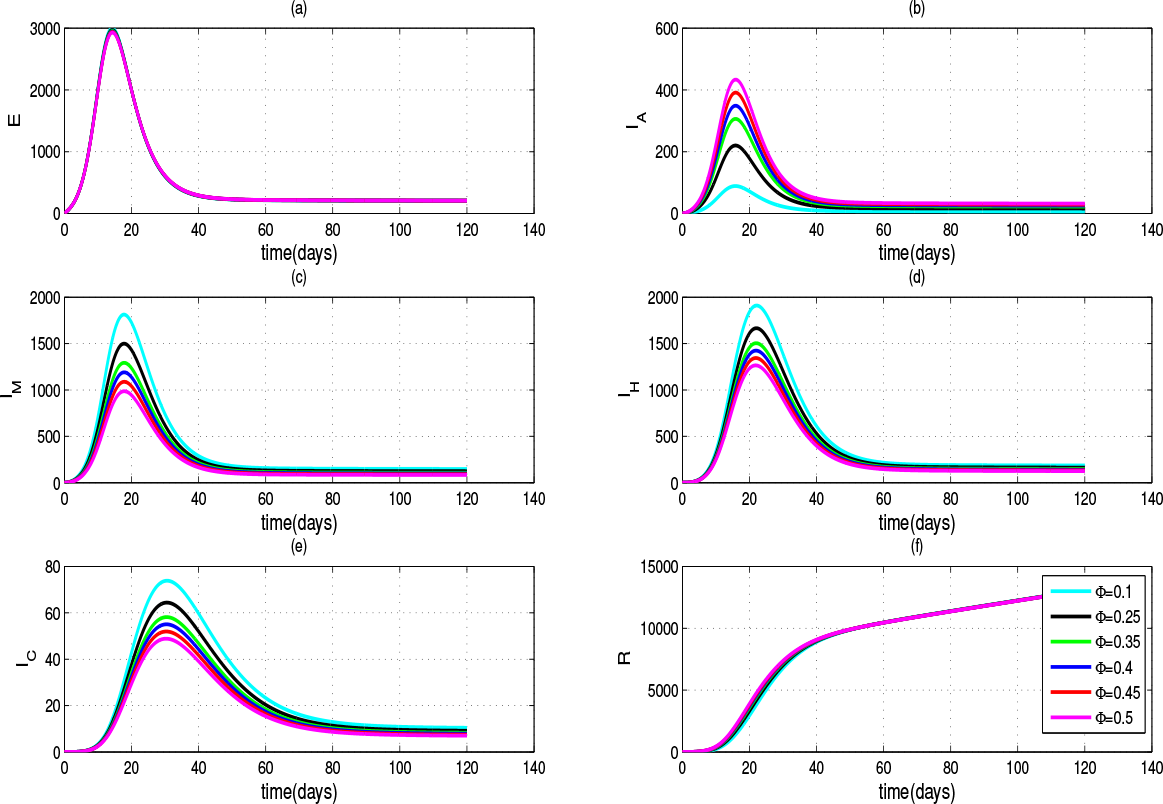
The simulated solution curves for the Exposed (a), Asymptomatic (b), Symptomatic with mild (c), Isolated in hospitals (d), Treated in ICUs (e) and Recovered (f) as given in (1) with varying parameter *φ* = (0.1, 0.25, 0.35, 0.4, 0.45, 0.5).

It is very clearly seen from Figure 3 that as *φ* increases, the number of asymptomatic cases also increase and this critical early diagnostic strategy has helped number of hospitalizations (*I_H_*) and that of severely sick patients (*I_C_*) to reduce.

Solution trajectories of Exposed *E* population onto Asymptomatic *I_A_*, Symptomatic with mild *I_M_*, Isolated in hospitals *I_H_* and Critically sick *I_C_* are presented respectively in Figure 4 (a)-(d).

**Fig. 4:**
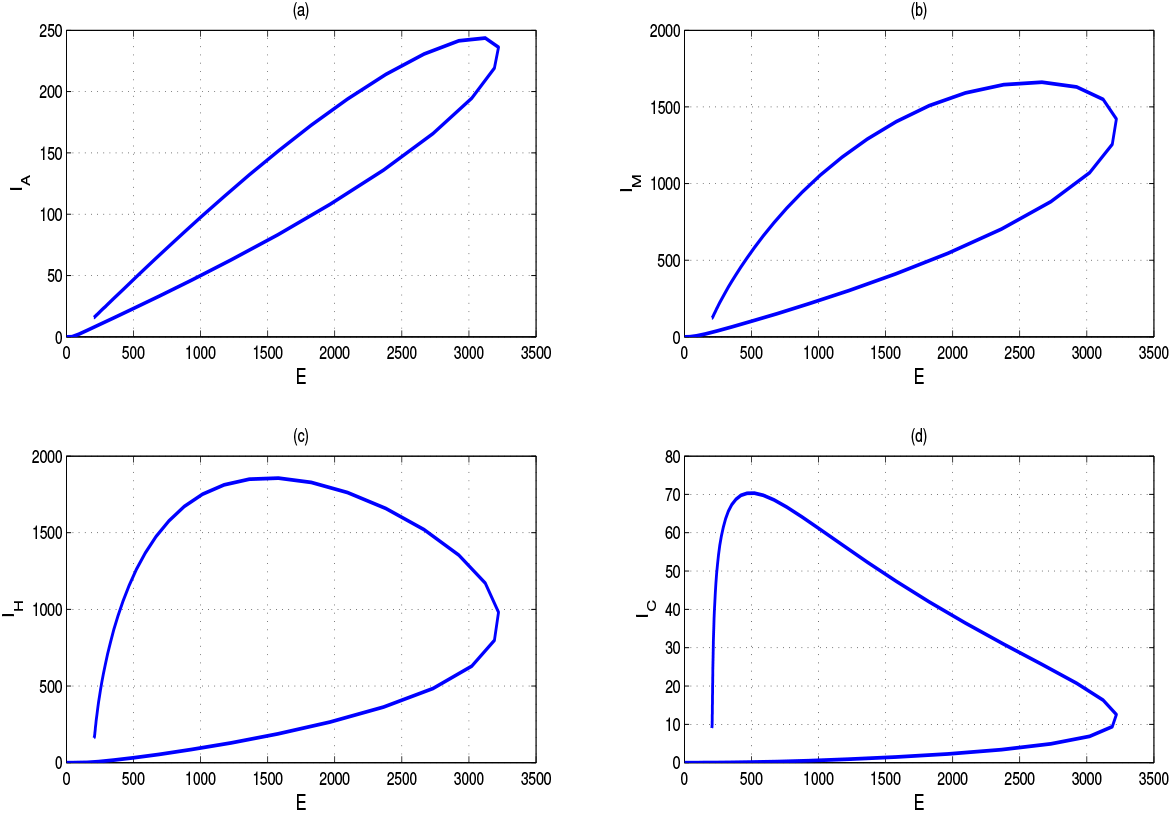
Solution trajectories (*E, I_A_*), (*E, I_M_*), (*E, I_H_*) and (*E, I_C_*) for the problem 1 with fixed parameter *φ* = 0.25.

### 4.3 Simulation of the Control Problem

In this section, we evaluate the efficacy of our three control measures, personal protection and social distancing (*u*_1_ ≠ = 0), diagnostic and isolation of asymptomatic cases (*u*_2_ ≠ = 0) and diagnostic and isolation of mild symptomatic cases (*u*_3_ ≠ = 0). First we simulate the problem in (10) considering non optimal control measures. We consider three combinations (*u*_1_ = 0.75*, u*_2_ = 0.5*, u*_3_ = 0.5), (*u*_1_ = 0.5*, u*_2_ = 0.3*, u*_3_ = 0.3) and (*u*_1_ = 0.25*, u*_2_ = 0.2*, u*_3_ = 0.2). The simulated results are given in Figure 5. According to Figure 5, it is clearly seen that when the control measures are increased the curves are flatten and the peak is occurred with a delay so that the public health system and hospitals can be prepared to handle the outbreak.

**Fig. 5:**
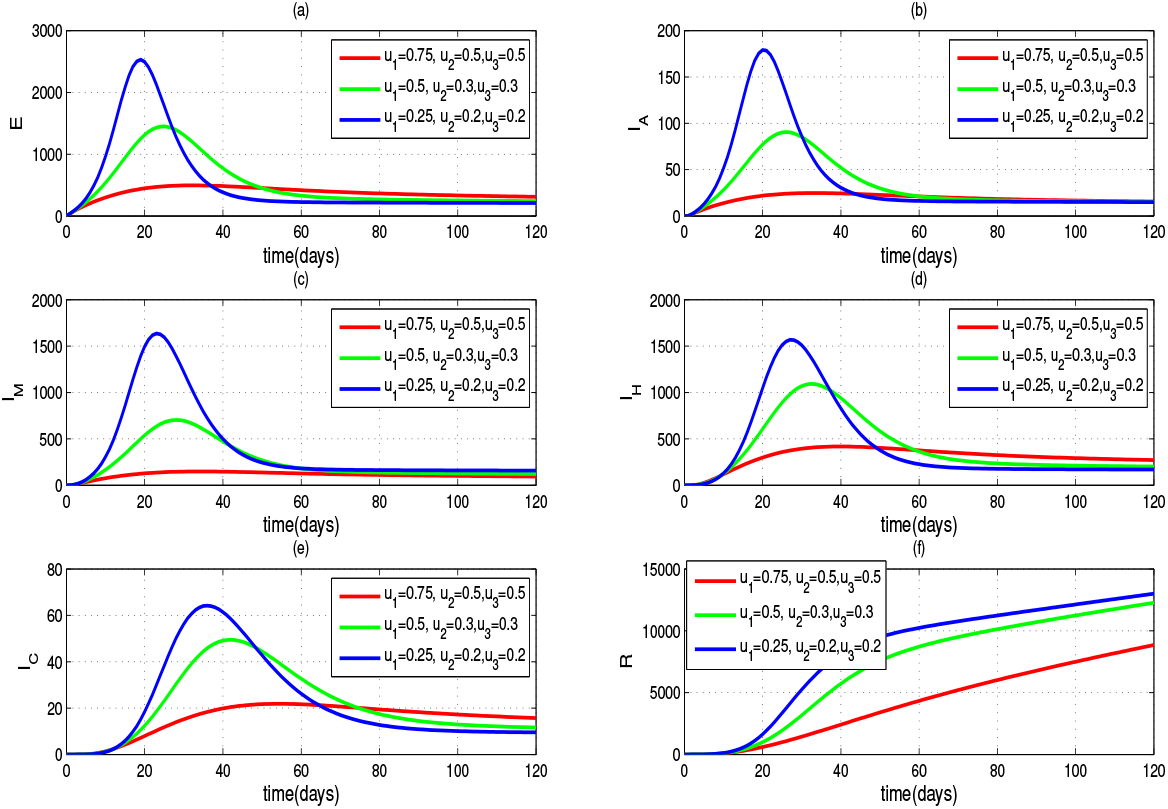
The simulated solution curves for the Exposed (a), Asymptomatic (b), Symptomatic with mild (c), Isolated in hospitals (d), Treated in ICUs (e) and Recovered (f) as given in problem (10) considering combinations of non optimal control measures (*u*_1_ = 0.75*, u*_2_ = 0.5*, u*_3_ = 0.5), (*u*_1_ = 0.5*, u*_2_ = 0.3*, u*_3_ = 0.3) and (*u*_1_ = 0.25*, u*_2_ = 0.2*, u*_3_ = 0.2).

The cost functional given in (12) is used to compute the associate cost for the government if non optimal control measures are introduced. The cost incurred if u_1_ = 0.75, u_2_ = 0.5, u_3_ = 0.5 is 4.9214 × 10^6^, if u_1_ = 0.5, u_2_ = 0.3, u_3_ = 0.3 is 4.0192 × 10^6^, and if u_1_ = 0.25, u_2_ = 0.2,u_3_ = 0.2 is 3:6519 × 10^6^.

The main goal of the optimal control problem presented in (10)-(20) is to minimize the number of exposed (*E*), asymptomatic infected cases (*I_A_*) and mild symptomatic infected cases (*I_M_*). In the public health point of view, it is aimed to reduce the number of patients who are in the community and able to transmit the virus, and make them isolated in designated hospitals. The simulation of the optimal control problem (10)-(20) is performed over three scenarios based on the relative importance of the three control measures. The parameters are *β*_1_ = 0.5*, β*_2_ = 0.6*, β*_3_ = 0.45*, γ*_1_ = 0.5*, γ*_2_ = 0.2*, γ*_3_ = 0.05*, η* = 0.005*, µ* = 0.04*, σ* = 1*/*5*, φ* = 0.25, *k* = 0.00405 and *q* = 0.0004. The initial conditions for the problem are *S*(0) = 0.85*, E*(0) = 0*, I_A_*(0) = 0*, I_M_*(0) = 0*, I_H_*(0) = 0*, I_C_*(0) = 0 and *R*(0) = 0.

#### 4.3.1 Scenario 1

We assume the social distancing and personal protection measures are highly important while the costs on two diagnostic and isolation are equal. The simulated outcomes for each populations *E, I_A_, I_M_, I_H_, I_C_* and *R* are presented in Figure 6 while the time invariant functions *u*_1_(*t*), *u*_2_(*t*) and *u*_3_(*t*) are illustrated in Figure 7.

**Fig. 6:**
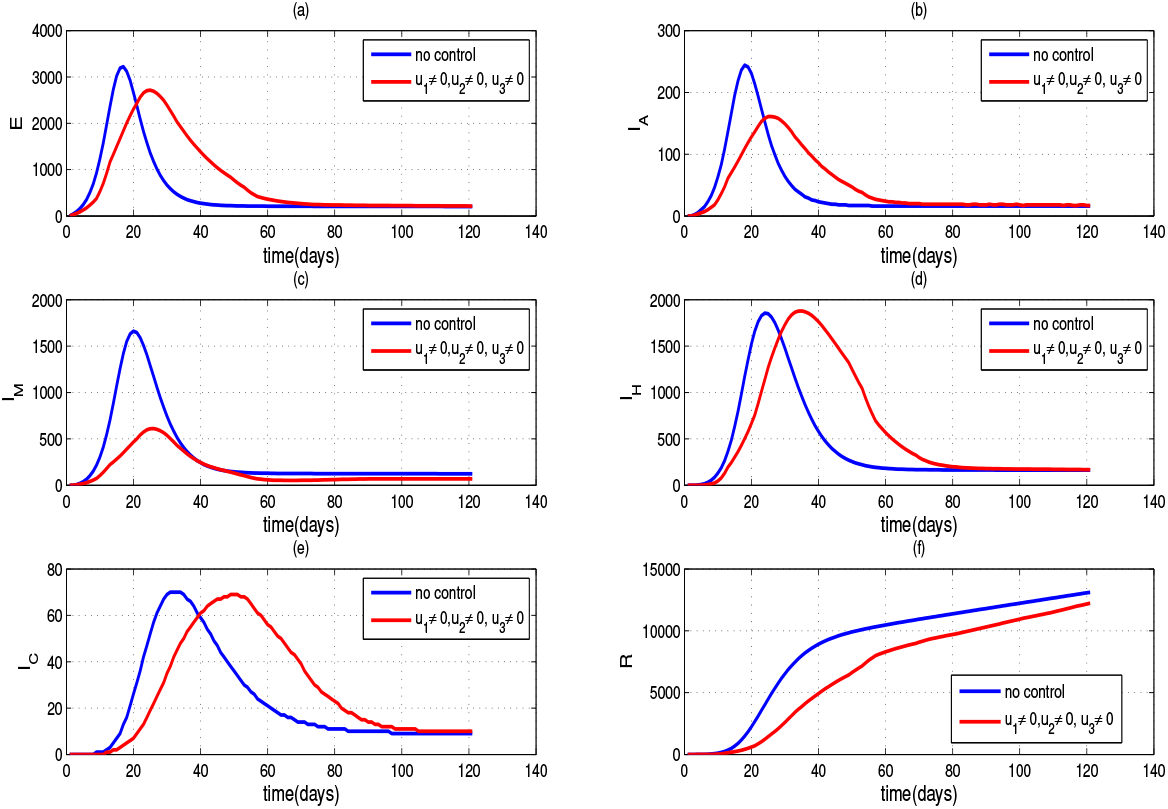
The simulated solution curves for the Exposed (a), Asymptomatic (b), Symptomatic with mild (c), Isolated in hospitals (d), Treated in ICUs (e) and Recovered (f) for the optimal control problem given in (10)-(20) with *A*_1_ = 50, *A*_2_ = 75, *A*_3_ = 60, *C*_1_ = 8, *C*_2_ = *C*_3_ = 2. It is assumed that the relative cost for social distancing and personal protection is high.

**Fig. 7:**
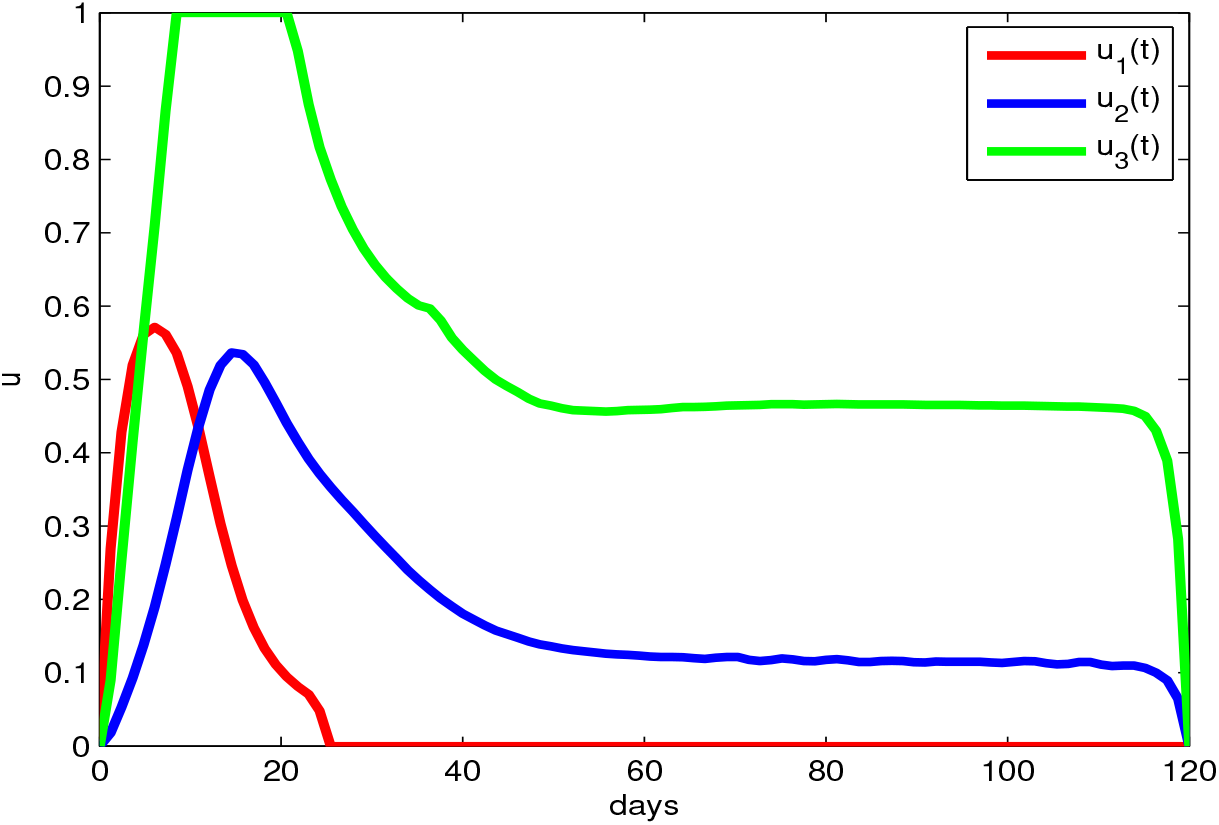
The optimal control profiles *u*_1_(*t*), *u*_2_(*t*) and *u*_3_(*t*) with *A*_1_ = 50, *A*_2_ = 75, *A*_3_ = 60, *C*_1_ = 8, *C*_2_ = *C*_3_ = 2.

It is seen from the Figure 6 that the control interventions are effective since the number of cases for each *E, I_A_* and *I_M_* populations have reduced compared to they are for the problem with out control in (1). Further, it is seen that the peak of each curve has been reduced and it is delayed. Thus, the optimal control measures have helped to flatten the curve. The control functions in Figure 7 suggest that tracing, testing and isolation of both asymptomatic and symptomatic infections are required for the entire period of time considered for the simulation.

#### 4.3.2 Scenario 2

We assume that tracing, testing and isolating asymptomatic cases are more critical. The simulated outcomes for each populations *E, I_A_, I_M_, I_H_, I_C_* and *R* are presented in Figure 8 while the time invariant functions *u*_1_(*t*), *u*_2_(*t*) and *u*_3_(*t*) are illustrated in Figure 9.

**Fig. 8:**
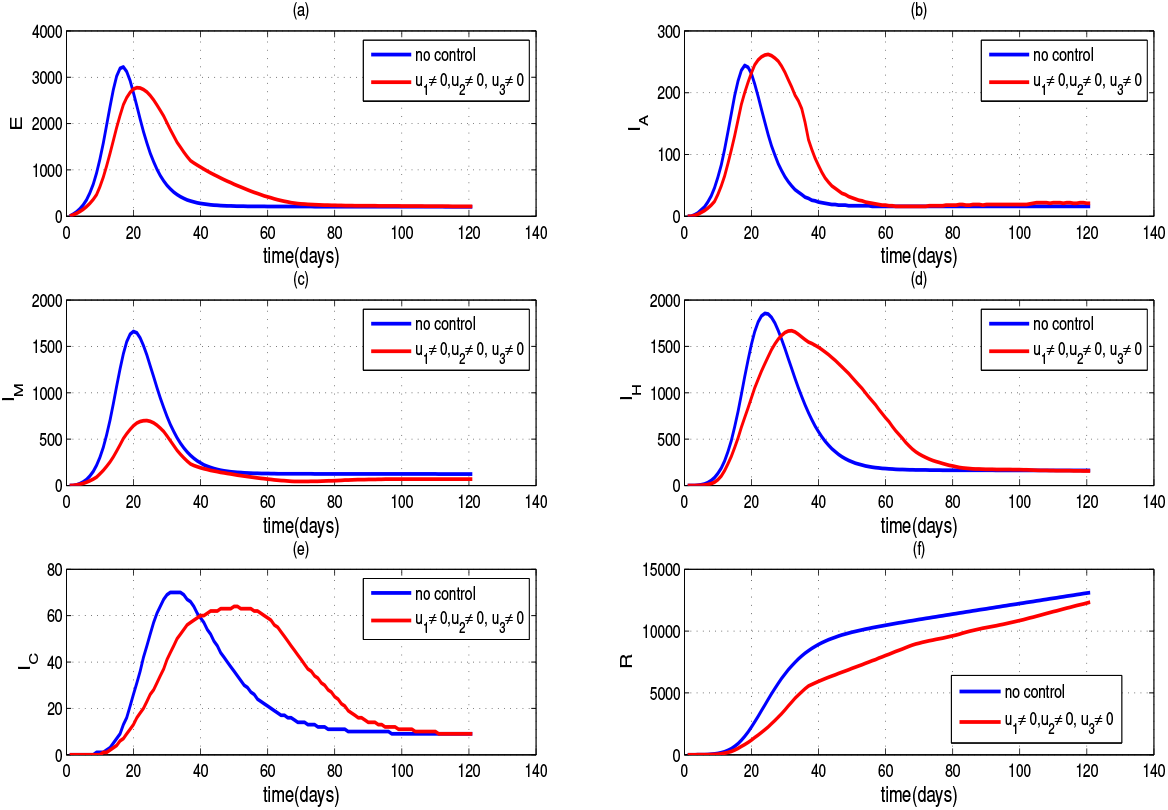
The simulated solution curves for the Exposed (a), Asymptomatic (b), Symptomatic with mild (c), Isolated in hospitals (d), Treated in ICUs (e) and Recovered (f) for the optimal control problem given in (10)-(20) with *A*_1_ = 50, *A*_2_ = 75, *A*_3_ = 60, *C*_1_ = 5, *C*_2_ = 8,and *C*_3_ = 2. It is assumed that the relative cost for tracing and testing asymptomatic cases is high.

**Fig. 9:**
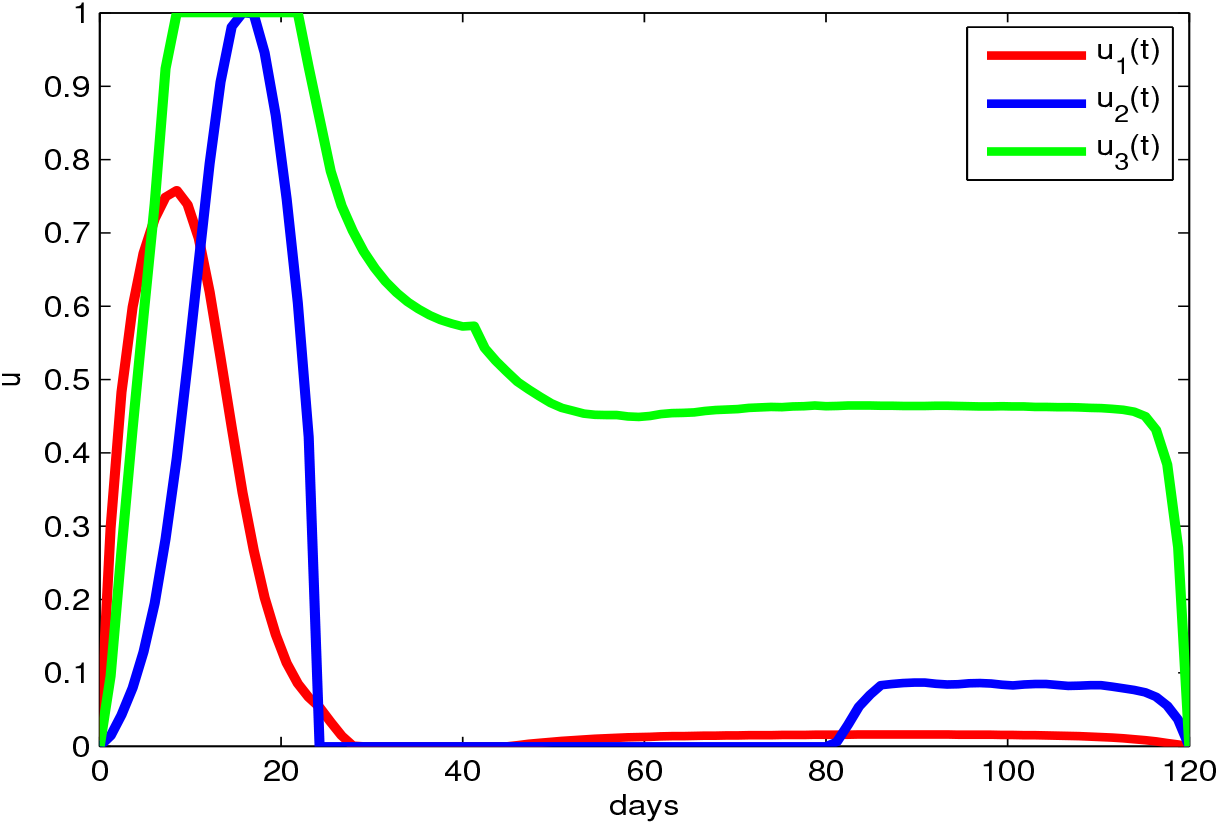
The optimal control profiles *u*_1_(*t*), *u*_2_(*t*) and *u*_3_(*t*) with *A*_1_ = 50, *A*_2_ = 75, *A*_3_ = 60, *C*_1_ = 5, *C*_2_ = 8, *C*_3_ = 2.

It is also seen from the Figure 6 that the control interventions are effective since the number of cases for each *E, I_M_, I_H_, I_C_* populations have reduced compared to they are for the problem with out control. All three control interventions needed in their full capacity during the initial stage of the outbreak, according toFigure 9.

#### 4.3.3 Scenario 3

We assume that social distancing with personal protection and tracing, testing and isolating mild asymptomatic cases are equally more critical. The simulated outcomes for each populations *E, I_A_, I_M_, I_H_, I_C_* and *R* are presented in Figure 10 while the time invariant functions *u*_1_(*t*), *u*_2_(*t*) and *u*_3_(*t*) are illustrated in Figure 11.

**Fig. 10:**
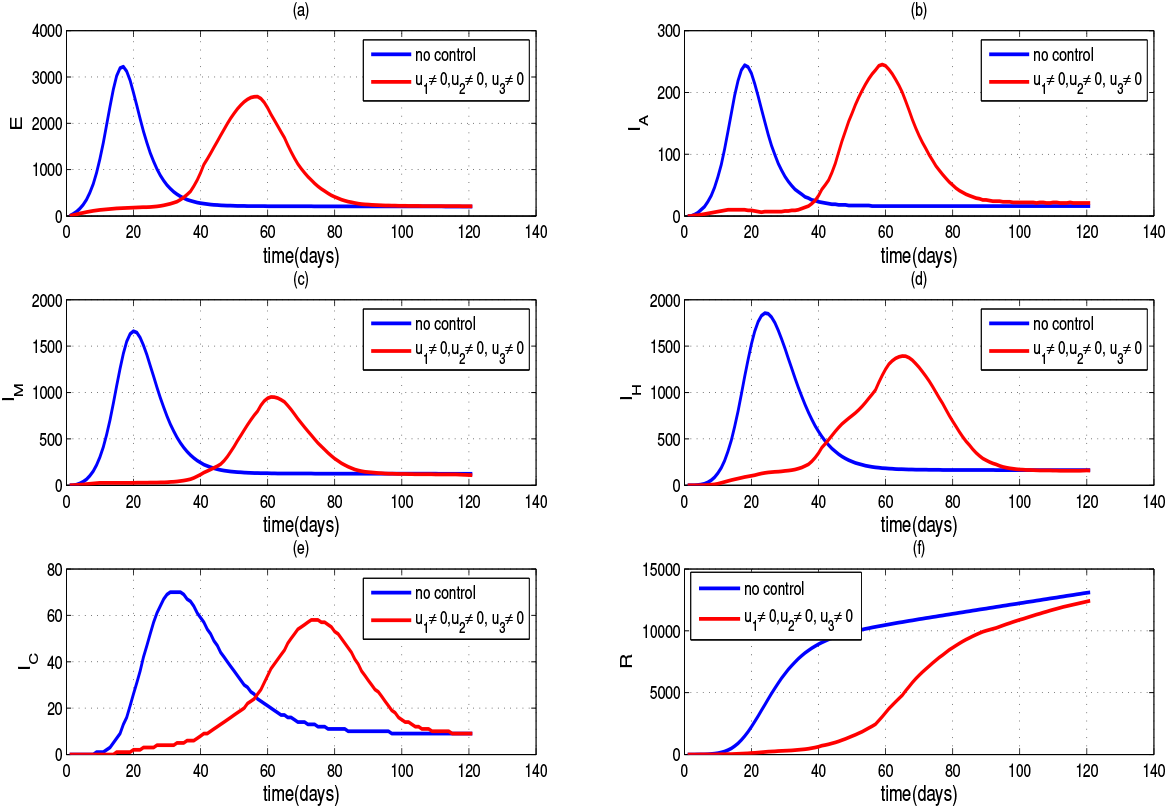
The simulated solution curves for the Exposed (a), Asymptomatic (b), Symptomatic with mild (c), Isolated in hospitals (d), Treated in ICUs (e) and Recovered (f) for the optimal control problem given in (10)-(20) with *A*_1_ = 50, *A*_2_ = 75, *A*_3_ = 60, *C*_1_ = 9, *C*_2_ = 9,and *C*_3_ = 3. It is assumed that the relative cost for tracing and testing symptomatic cases is high while less importance is given for social distancing and personal protection.

**Fig. 11:**
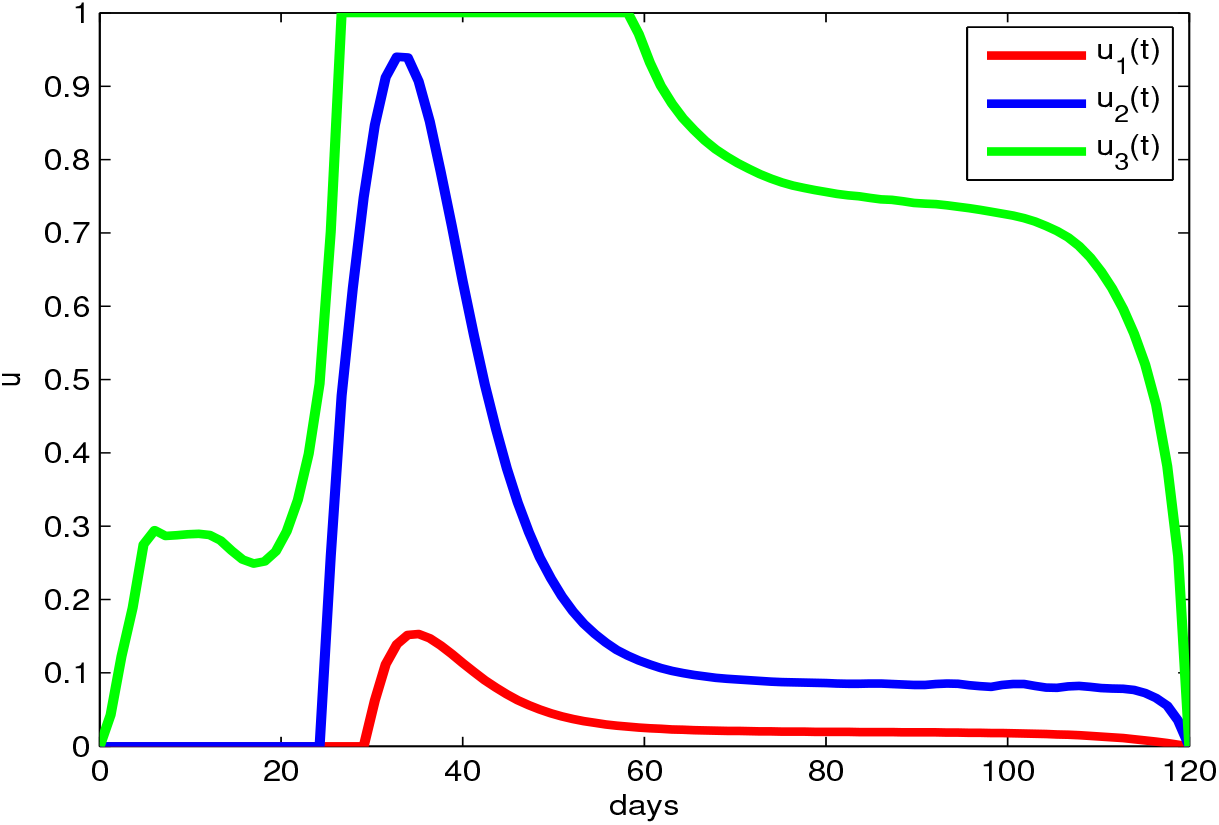
The optimal control profiles *u*_1_(*t*), *u*_2_(*t*) and *u*_3_(*t*) with *A*_1_ = 50, *A*_2_ = 75, *A*_3_ = 60, *C*_1_ = 9, *C*_2_ = 9, *C*_3_ = 3.

According to Figure 10, if the health system focuses equally more on social distancing and personal protection, tracing of asymptomatic cases then the peak of the exposed, asymptomatic, symptomatic, hospitalized, and ICU treated cases can be minimized on the other hand each peak can be delayed. Therefore, it can be stated, this control strategy is successful as the government needs to encourage more on social distancing and personal protection practices together with effective tracing, testing and isolation strategy for the patients who do not show symptoms.

The algorithm for the optimal control problem was iterated 100 times until the optimal solutions are found. The cost functional given in (12) is evaluated in each iteration and the behavior of execution is given in Figure 12.

**Fig. 12:**
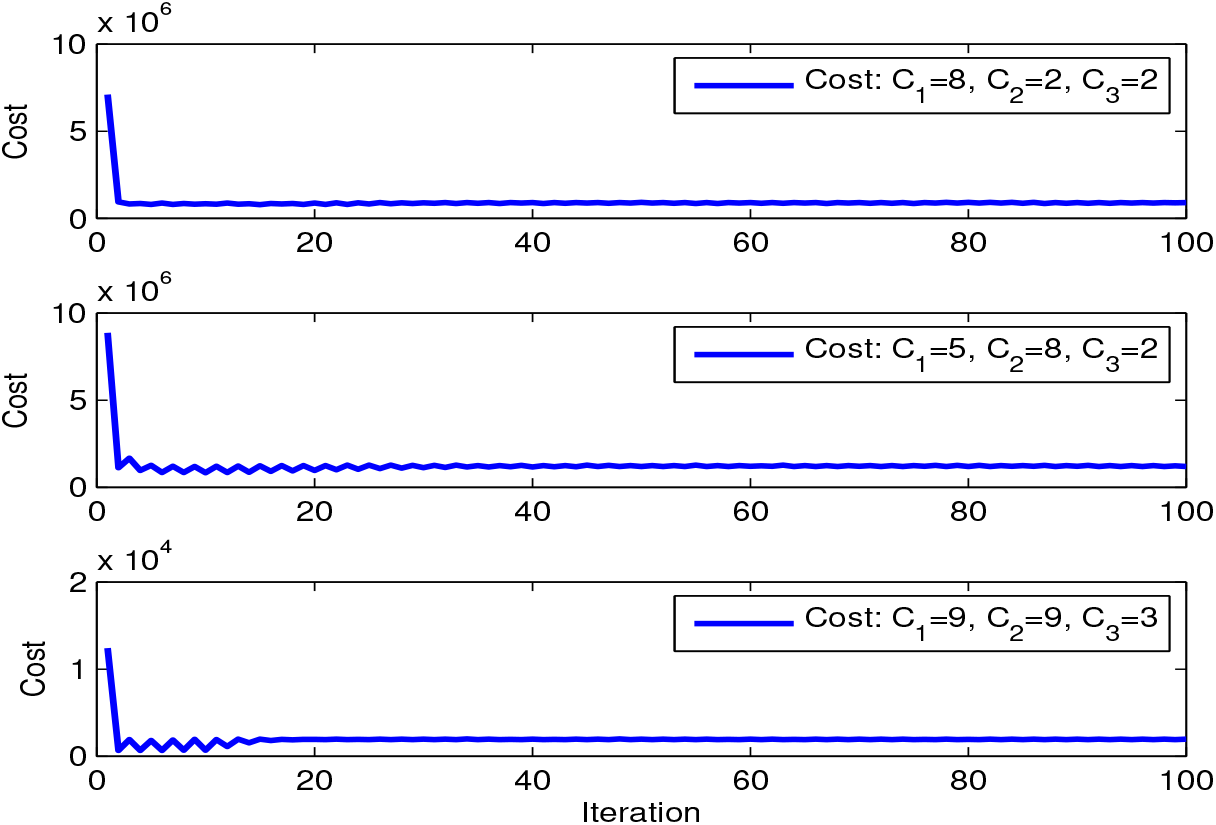
The behavior of cost functional given in (12) with respect to iterations.

It can be clearly seen the convergence of cost to its optimal value 9:035 × 10^5^ units for the scenario 1 while it is for scenario 2 obtained as 11:88 × 10^5^ units, however, for the scenario 3, the cost is as small as 1:95 × 10^3^.

## 5 Conclusion

Presently, COVID 19 disease caused by the novel corona virus has been a very serious public health concern across the globe. This outbreak is more than five months old since it was first claimed to be originated in the city of Wuhan, China in late December 2019. The development of suitable vaccine candidate is still in progress thus, strong social control measures and public health interventions are critically needed to combat with the disease spread.

COVID 19 is a novel disease, therefore researchers are learning about the dynamic of this virus everyday. In an epidemiological state in this type, mathematical models are very useful to understand the dynamic of the disease and to evaluate the efficacy of different control measures such as social distancing, personal protection, and public health interventions such as contact tracing, isolation and treatments.

In this study, we develop an extended version of SIER conceptual model considering two main clinical, epidemiological and public health facts; firstly, the occurrence of asymptomatic and symptomatic infections of people and secondly, the individual demography such as age, life style and health condition found to have determined the patient’s situation might turn into severe.

Since the government works hard utilizing most of its resources to control the spread over the population, an optimal control model is also constructed. Essential mathematical analysis is carried out for the models to check the stability of the equilibrium points, derive disease’s *R*_0_, investigate the existence of solutions to the optimal control problem, and etc while numerical simulations are performed in MATLAB. It is clearly seen from the simulations presented in Figures 7–11 that optimal control measures have reduced the exposed, asymptomatic, symptomatic cases significantly. The control scenario 3 provides a considerable effect on the epidemic curves, not only it minimizes the infections but also delaying the peak of the outbreak approximately by 40 days in contrast to the outcomes with out control. This enables the health system to be more equipped and prepared to combat with the epidemic. It should be noted that the simulations are carried out for a period as short as for 120 days.

## Data Availability

This is a mathematical model and computer simulations generated data. No real data used.

